# In Vivo Positronium Lifetime Measurements with Intravenous Tracer Administration and a Long Axial Field-of-View PET/CT

**DOI:** 10.1101/2024.10.19.24315509

**Authors:** Lorenzo Mercolli, William M. Steinberger, Hasan Sari, Ali Afshar-Oromieh, Federico Caobelli, Maurizio Conti, Ângelo R. Felgosa Cardoso, Clemens Mingels, Paweł Moskal, Thomas Pyka, Narendra Rathod, Robin Schepers, Ewa Ł. Stępień, Marco Viscione, Axel Rominger, Kuangyu Shi, Robert Seifert

## Abstract

**Purpose:** Measuring orthopositronium (oPs) lifetime could in principle provide diagnostic information about the tissue microenvironment that goes beyond standard positron emission tomography (PET) imaging. This study demonstrates that in vivo oPs lifetime measurement is feasible with a commercial long axial field-of-view (LAFOV) PET/CT scanner.

**Methods:** Three subjects received a dose of 148.8 MBq [^68^Ga]-Ga-DOTA-TOC, 159.7 MBq [^68^Ga]Ga-PSMA-11 and 420.7 MBq [^82^Rb]Cl. In addition to the standard protocol, the three subjects were scanned for 20, 40 and 10 minutes with a single-crystal interaction acquisition mode on a Biograph Vision Quadra (Siemens Healthineers) PET/CT. Three-photon events, that include two annihilation photons and a prompt photon from the decay of the radionuclide, are then selected and localized with time-of-flight using a prototype software. Through a Bayesian fit to the measured time difference between the annihilation and the prompt photons, we are able to determine the oPs lifetime for selected organs. The Bayesian fitting methodology is extended to a hierarchical model in order to investigate possible common oPs lifetime distributions of the heart chambers in the [^82^Rb]Cl scan.

**Results:** From the segmentation of the subjects’ histoimages of three-photon events, we present the highest density intervals (HDI) of the oPs lifetime’s marginalized posterior distribution for selected organs. Interestingly, the mean values of the right heart chambers were higher than in the left heart chambers of the subject that received [^82^Rb]Cl: the 68% HDI of the atria are [1.15 ns, 1.72 ns] (left) and [1.46 ns, 1.99 ns] (right) with mean values 1.50 ns and 1.76 ns, respectively. For the ventricles we obtained [1.22 ns, 1.60 ns] (left) and [1.69 ns, 2.18 ns] (right) with mean values 1.44 ns and 1.96 ns. This might signal the different oxygenation levels of venous and arterial blood. Fitting a hierarchical model, we found that the oPs lifetime for volumes-of-interest with arterial blood can be sampled from a posterior distribution with a 68% HDI of [1.4 ns, 1.84 ns] (mean 1.62 ns) and while those containing venous blood have a HDI of [1.78 ns, 2.21 ns] (mean 2.0 ns). The ^68^Ga measurements showed significantly worse count statistics and therefore larger errors in the oPs lifetime.

**Conclusion:** In vivo oPs lifetime measurements on a commercial LAFOV PET/CT system are feasible at the organ level with an unprecedented level of statistical power. Nevertheless, count statistics of three-photon events and the interpretation of oPs lifetimes in human tissue remain major challenges that need to be addressed in future studies.

## I. ntroduction

**P**OSITRONIUM (Ps), the bound state of an electron and a positron, has been an object of investigation for more than a century. The spin singlet state is called parapositronium (pPs) and the triplet state orthopositronium (oPs). The distinctive feature of Ps compared to a hydrogen atom is the possibility to annihilate. pPs and oPs are odd and even eigenstates of charge conjugation and parity transformations. The corresponding selection rule therefore implies that the dominant annihilation mode for pPs is in two photons, while oPs will annihilate into three photons. Even using the most naïve Ps decay model, the decay constant of oPs in vacuum is almost three orders of magnitude times smaller than the decay constant of pPs

Being relative long-lived, oPs has enough time undergo interactions with surrounding material. The so-called *pick-off* process describes the oPs annihilation to two *γ* from an interaction with an environmental electron (see e.g. Refs. [1]–[3]). Such a pick-off shortens the oPs lifetime significantly and introduces a dependence on the physical and chemical environment. Apart from the pick-off process, spin exchange with unpaird electrons can lead to a conversion from oPs to pPs as well and thus a two fast *γ* annihilation as well as oxidation [1], [3]. The oPs annihilation into two photons therefore becomes a surrogate quantity for the molecular structure of the surrounding material. It should be noted that the probability of Ps formation also depends on environmental conditions.

The measurement of oPs lifetime is often termed *positron lifetime spectroscopy* (PALS). It can provide detailed information about defects in solids, lattice imperfections in semiconductors, free volumes and structures in polymers (see e.g. Refs. [1]–[5]). The high sensitivity to a material’s structure and relatively simple experimental setups made PALS a widely adopted technique in material science.

The medical domain has shown significant interest in PALS (see e.g. Refs. [2], [3], [6]–[14]). Driven by the possibility to measure oxygenation levels in human tissue [15]–[20], several in vitro studies were performed by the authors of Refs. [7], [11], [21]. Recently, the first in vivo measurement of oPs lifetimes was reported in Ref. [22]. The oPs lifetime has the potential to add diagnostic information which is currently unavailable or requires additional interventions, such as e.g. biopsy or additional use of hypoxia tracers.

A key factor in exploring the clinical potential of oPs lifetime measurements is the ability to use standard positron emission tomography (PET) scanners and a clinically routine way of intra venous tracer administration [23]. Standard PET/CT scanners are devised and optimized to detect two coinciding 511 keV photons from the positron annihilation. However, the oPs lifetime is determined through the measurement of the time difference between the detection of two annihilation photons and a prompt photon from the decay of the mother nuclide. This means there is a need to identify three-photon events (3*γ*E) rather that just two-photon coincidences (and that such measurements can be done only with non-pure positron emitters [24]). Therefore, the detection, storage and processing of single photon interactions in the PET detector are mandatory to measure the oPs lifetime, as introduced e.g. in Ref. [25] for the J-PET detector with the trigger-less data acquisition.

Another major challenge in PALS measurement is the collection of enough 3*γ*E. Typical PALS measurement use long-lived radionuclides with a high prompt gamma branching fraction, like e.g. ^22^Na, and data collection can last for many hours. However, radionuclides used in clinical routine have only a limited prompt photon branching fraction, if any at all, and scan time is limited by either a short half-life, as in the case of ^82^Rb, or by the time a patient is able to stay in the scanner. This is why only long axial field-of-view (LAFOV) PET/CT systems stand a chance to collect the necessary count statistics for in-vivo Ps lifetime measurements. This kind of systems has entered clinical routine operation a couple of years ago (see e.g. Refs. [26]) and has shown to have a significant increase in sensitivity over standard axial field-of-view scanners [27], [28]. Note that also the image reconstruction process for 3*γ*E has triggered substantial research, as shown e.g. in Refs. [29]–[34], in order to address the problem of limited count statistics.

Recently, we showed in Refs. [35] that it is possible to determine the oPs lifetime in reference materials with a Siemens Biograph Vision Quadra (Siemens Healthineers, Knoxville, TN, USA) (henceforth simply referred to as Quadra) with the clinically used radionuclides ^68^Ga, ^124^I and ^82^Rb. Quadra has a prototype feature that allows for a trigger-less acquisition mode, the so called *singles mode*, where single-crystal interactions within a given mini-block detector are saved without coincidence sorting to a list mode file. However, the feasibility of in vivo oPs lifetime measurements with an intravenously administered tracer as done in routine clinical practice and a clinical LAFOV PET/CT system is currently not known.

Therefore, in the present manuscript, the first in vivo Ps lifetime measurements with a commercial LAFOV PET/CT system and a routine route of tracer administration is proposed. Using the event selection algorithm presented in Ref. [35], we determine the time difference spectra for different organs. Through a Bayesian fitting procedure we were then able to estimate the oPs lifetime for those organs. Finally, within the Bayesian framework, we developed a hierarchical model to combine 3*γ*E from different volume-of-interest (VOI).

## II. Materials and methods

### A. Study subjects and scan protocols

This study included two patients and a healthy volunteer. The personal details are given in Tab. I. The subjects were administered with different compounds with ^68^Ga and ^82^Rb. Both radionuclides emit prompt photons, but with a relatively low BR of 1.190 ± 0.017 for ^68^Ga and 13.5 ± 0.5 for ^82^Rb per positron emission (see e.g. https://www-nds.iaea.org/).

**TABLE I:** Details of the three subjects and administered activities.

| Subject | Weight [kg] | Radiopharm. | Activity [MBq] |
| --- | --- | --- | --- |
| S1 | 71 | $^{68}\text{Ga}$]Ga-DOTA-TOC | 148.8 |
| S2 | 78 | $^{68}\text{Ga}$]Ga-PSMA-11 | 159.7 |
| S3 | 80 | $^{82}\text{Rb}$]Cl | 420.7 |

A woman (referred to as S1 in the following) with multifocal meningioma was administered 148.8 MBq of [^68^Ga]Ga-DOTA-TOC. A 10 min acquisition in coincidence mode, i.e. a standard clinical scan with two-photon coincidence sorting, was performed 32 min p.i. (see Tab. II). At 44 min p.i. the singles mode scan was started and lasted 20 min. Fig. 1 shows the coronal and sagittal maximum intensity projections (MIP) of the coincidence PET image.

**TABLE II:** Start time and duration in minutes of the singles and coincidence PET scans.

| Subject | Coin. | Start coin. | Singles | Start sing. |
| --- | --- | --- | --- | --- |
| S1 | 10 | 32 p.i. | 20 | 44 p.i. |
| S2 | 13 | 123 p.i. | 40 | 18 p.i. |
| S3 | - | - | 10 | 0 p.i. |

**Fig. 1:**
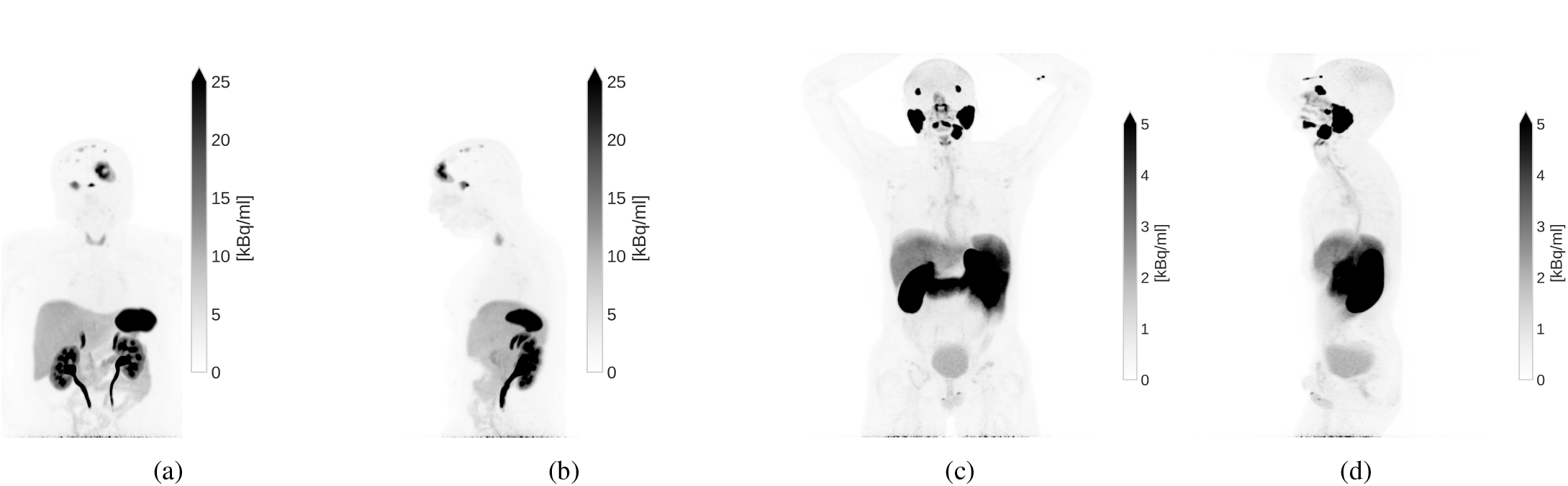
Coronal and sagittal MIP of the coincidence PET images for S1 (1a and 1b) and S2 (1c and 1d).

The second patient S2 was a man with a prostatic adenocarzinoma and a suspected local recurrence. He had undergone a prostatectomy about three years prior to the this study. The patient received 159.7 MBq [^68^Ga]Ga-PSMA-11 and was scanned for 13 min in coincidence mode about two hours p.i. As summarized in Tab. II, the singles mode scan lasted for 40 min and started prior to the clinical PET/CT scan. The early coincidence PET image is shown in Fig. 1.

Finally, the healthy volunteer S3 obtained 420.7 MBq [^82^Rb]Cl. The [^82^Rb]Cl was produced with CardioGen-82 radionuclide generator and infusion system (Bracco Imaging S.p.A, Milan, Italy). Due to the short half-life of ^82^Rb, only the singles mode acquisition and a low-dose CT scan were performed for S3. The infusion of the [^82^Rb]Cl started simultaneously with the singles mode acquisition and lasted for 19 s.

We evaluate several organs for oPs lifetime measurement across all three subjects, including the kidneys, liver, lungs, pancreas, and spleen. In addition, specific organs are segmented for each subject: S1 had the meningioma segmented; S2 included the parotid gland. The suspected local recurrence in S2 is too small to provide a sufficient number of 3*γ*E for analysis. For S3, the additional VOI are the aorta, right axillary vein, brain, thyroid, and the four heart chambers. Most segmentations are performed on CT images using the AI-based TotalSegmentator tool [36]. An experienced nuclear medicine clinician reviewed and amended the automatic segmentations wherever needed. The meningioma in S1 is segmented using the coincidence PET image and the axillary vein in S3 is directly segmented using the histoimage of 3*γ*E (image with number of detected 3*γ*E as voxel value). In particular for the heart chambers, aorta and axillary vein, the VOIs were segmented conservatively to mitigate partial volume effects due to inaccurate localization of 3*γ*E.

### B. Data acquisition

The details of the 3*γ*E selection and time difference measurements with Quadra are discussed in Refs. [35]. In singles mode, Quadra registers all detector hits without any triggering or event sorting and writes the photon energy, time stamp and crystal ID into a list-mode file. A prototype software groups all possible combinations of three detector events within a time window of 20 ns into a 3*γ*E. In a second step, all 3*γ*E that do not fall within a given energy window are discarded. Two energy windows need to be selected: one for the two annihilation photons and one for the prompt photon. For both ^68^Ga and ^82^Rb, we set the energy window for the annihilation photons from 435 to 585 keV and the prompt photon window from 720 to 726 keV. As explained in Ref. [35], the last energy bin collects all detector hits from higher energies. I.e. the prompt photons of ^68^Ga and ^82^Rb are detected in the last energy bin, but their energy is not resolved. Next, the two annihilation photons must be detected within a coincidence time window of 4.2 ns for the 3*γ*E not to be discarded. Finally, a threshold on the minimum distance between the detection location of the prompt and annihilation photons is enforced. Due to the spatial correlation of the detector hits originating from the ^176^Lu background in Quadra’s LSO crystals, this spatial threshold effectively removes false 3*γ*E from crystal radiation (see Ref. [35] for details). No additional restriction on the acceptance angle is imposed compared to standard coincidence acquisition mode.

Once the 3*γ*E are sorted, the spatial location of the 3*γ*E is reconstructed with the time-of-flight information of the two annihilation photons. From the 3*γ*E location and the flight path of the prompt photon, it is then straight forward to determine the time difference between the prompt photon emission and positron annihilation. The histograms of the measured time differences for all 3*γ*E in a VOI are the time difference distributions (TDD). We do not account for the attenuation of photons in the subject’s bodies. Rescaling the number of detected photons by an attenuation map would likely rescale a TDD but not significantly change its shape.

### C. Data analysis

After the positron is emitted from the decaying nucleus, it undergoes many different processes and states before annihilating with an electron. Therefore, measured TDD contains many different components that we have to disentangle. Depending on the time scale, it is possible to neglect some of these components.

At first, emitted positrons will loose their energies mostly through interactions with the surrounding atoms (Bremsstrahlung, ionizations and inelastic collisions). At low energies, the annihilation cross section is proportional to (1 + 𝒪 (*β*))*/β* for *β* = *v/c*, i.e. the relative velocity of the electron and positron normalized to the speed of light. This makes a direct in-flight annihilation unlikely. It takes a 1.5 MeV positron about 𝒪 (20 ps) to slow down to 𝒪 (10 eV) in water. This time scale is rather short and is typically neglected in PALS measurements. Its effect in TDD is typically absorbed in the modeling of the measurement device. The Ps formation cross section peaks around 10 eV.If no Ps formation occurred, the positron will thermalize through elastic interactions and eventually annihilate with an electron. The thermalization process can take 𝒪 (400 ps) and cannot be neglected. One component of the TDD is therefore the direct annihilation without Ps formation.

Once a Ps state is formed, there are multiple possibilities for an annihilation into two photons, as illustrated e.g. in Fig. 1 of Ref. [15]. On average, pPs annihilates into two photons within ≈ 2*/*(*α*^5^*m*) ≈125 ps without being particularly affected by the environment. oPs can either turn into pPs through spin exchange or annihilate into two photons through a pick-off process. Since all annihilations in two photons are detected, both Γ(pPs → *γγ*) and Γ(oPs + *e*^−^ → *γγ*) are contributing to the measured TDD. The annihilation of oPs into three photons is not contained in the TDD since we do not sort four-photon events. In order to determine the oPs lifetime from the TDD, we therefore need to consider three components: direct annihilation together with pPs and oPs decay.

Note that the time difference measurement outlined in Sec. II-B assumes that the prompt photon emission has a negligible delay compared to the positron emission.

Quadra has a finite time resolution that was modeled through a single Gaussian with a width *σ* and expectation value Δ, which is convoluted with the sum of three exponential functions. The oPs lifetime *τ*_3_ is therefore determined by fitting the following model to the TDD of a given VOI

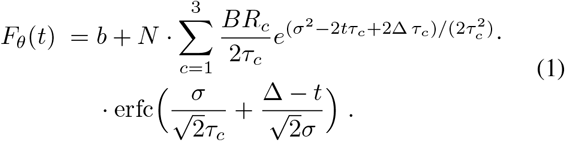

In Eq. (1), *b* denotes the background and *N* is a normalization constant. *BR*_*c*_ with *c* = 1 … 3 and ∑_*c*_ *BR*_*c*_ = 1 are the relative branching ratios of pPs, direct annihilation and oPs, respectively. The *θ* stands for the collection of all fitting parameters, i.e. *τ*_1,2,3_, *BR*_1,2,3_, Δ, *σ* and *N*. We only consider time differences from Δ*t*_0_ = − 1.5 ns to Δ*t*_*n*+1_ = 10 ns for the parameter fitting. We do not consider any additional lifetime components due to tissue inhomogeneities in a given VOI (e.g. in the kidneys). The oPs lifetime is therefore to be understood as an average value.

As in Ref. [35], we rely on a Bayesian framework for the fitting procedure. Given the challenging count statistics, the Bayesian fitting procedure allows us to accurately estimate the uncertainty of the oPs lifetime by marginalizing over the nuiscance parameters and to avoid common pitfalls of non-linear frequentist fits. Furthermore, within a frequentist framework we wouldn’t be able to construct and compare hierarchical models.

From the parameters in Eq. (1) we fix the background *b* to the median value of the TDD with time differences smaller than − 2 ns. All other parameters are sampled from prior distributions, which are mostly inspired by the values for water at 37^*°*^C reported in Ref. [37]. The priors for the lifetimes *τ*_*c*_ are

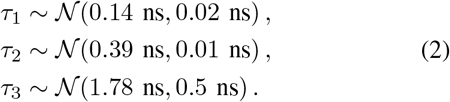

Note that the fitted value of *τ*_1_ from Ref. [37] differs from canonical *τ*_1_ = 125 ps (see e.g. Ref. [38]). This is likely due to the fact that the time it takes the positron to loose its energy prior to the pPs formation is not included in 125 ps. In any case, the prior of *τ*_1_ in Eq. (2) includes the value of 125 ps within less than one standard deviation. For *τ*_3_ we choose a rather wide and therefore uninformative prior distribution, which nevertheless accounts for our knowledge on the order of magnitude that we can expect in human tissue. The branching ratios of the three components follow a Dirichlet distribution, i.e.

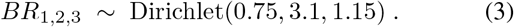

The mean values of the distribution in Eq. (3) are 0.15, 0.62 and 0.23 and the standard deviations are 0.15, 0.20 and 0.17. The knowledge of Quadra’s timing resolution allows us to set the prior distribution for *σ* and Δ to

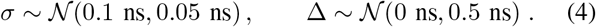

Finally, the normalization constant *N* is sampled from a normal prior distribution

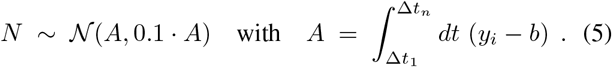

*y*_*i*_ with *i* = 1, …, *n* are all observations in a TDD, i.e. the number of counts in a time difference bin *i*. The use of the factor *A* in Eq. (5), which is purely determined by the measured TDD, significantly stabilizes the numerical calculation.

The count statistics are sufficiently high that we can assume a Gaussian likelihood. All observations *y*_*i*_ in a TDD are assumed to be independent and identically distributed (iid)

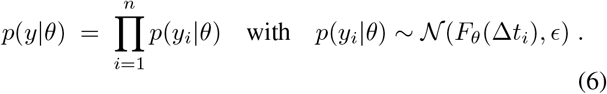

Δ*t*_*i*_ is the time difference of the bin *i* and the weakly informative prior on the variance *E*^2^ is the positive Cauchy-Lorentz distribution (see Ref. [39])

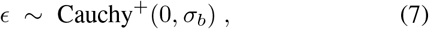

where 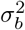 is the variance of the background *b* and is determined, analogously to *b* itself, as a point estimate from the measured TDD.

We report our results in terms of expectation values and 68% highest density intervals (HDI). In most cases, the posterior distributions are bell-shaped and the use of the standard estimator for the variance would be meaningful. However, this does not need to be the case, in particular for *BR*_*c*_, and we therefore stick to the HDI. The use of marginalized posterior distribution is particularly appealing since our main interest lies on the parameter *τ*_3_ and all others can in principle be considered as nuisance parameters.

The afore described Bayesian fitting procedure applies to the parameter fitting to a TDD from a single VOI. However, oPs lifetime could on average be the same for different VOI, if we have reason to believe that the tissue structure is similar. The example we explored in this study is the oPs lifetime in arterial and venous blood. Conceptually, this requires a comparison of statistical models with different number of fitting parameters. E.g. let us suppose there are two VOI that contain mostly venous blood. The question we seek to answer is whether a model where *τ*_3_ is sampled from one single distribution for the two VOI performs better than a model with two separate distributions for *τ*_3_. Therefore, we construct hierarchical models (see e.g. Ref. [40]) and rely on a leave-one-out cross-validation (LOO-CV) as discussed in e.g. Ref. [41].

Let us assume that we have *j* = 1, …, *n*_*j*_ different VOI. Each VOI has a TDD, which we shall denote as *y*_*ij*_ and a set of fitting parameters *θ*_*j*_. The likelihood *p*(*y*_*j*_ | *θ*_*j*_) is iid and factorizes as in Eq. (6). The fitting parameters *θ*_*j*_ are split into global parameters that are completely independent of the VOI and parameters that are VOI-dependent. *τ*_1,2_ are assumed to be independent of the tissue composition and the same applies to the scanner parameters *σ* and Δ. These parameters are sampled from a single prior distribution according Eqs. (2) and (4) and a sampled value is applied to all VOI. Next, the normalization *N*_*j*_ and statistical noise *E*_*j*_ are specific to every single VOI. They are sampled from separate prior distributions, but with the same prior parameters as in Eqs. (5) and (7). Finally, the main feature of the hierarchical model is that we can assume that the oPs lifetime and branching ratios are sampled from a common distribution for multiple VOI. Fig. 2 shows the basic scheme of Bayesian hierarchical models used in this study. The likelihood remains the same as in Eq. (6), simply with an additional group (or VOI) index *j*. However, the prior distribution of 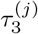and 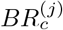 is conditional on a series of hyperparameters, which we denote as *ϕ*. Specifically,

**Fig. 2:**
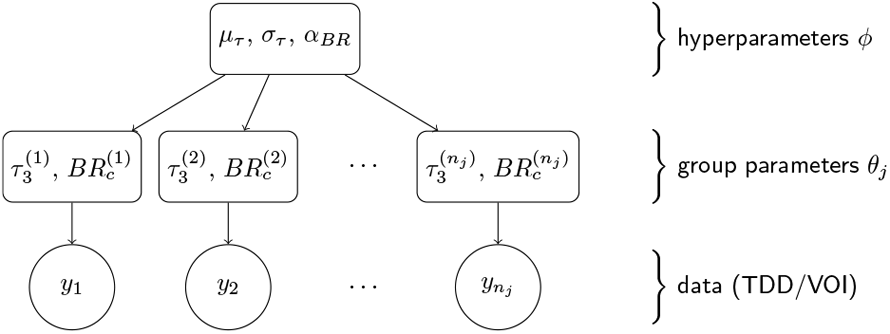
Hierarchical model for the combined analysis of data from different VOI.

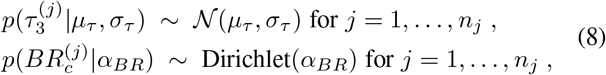

and the hyperparameters *µ*_*τ*_, *σ*_*τ*_, and *α*_*BR*_ are sampled from the following prior distributions

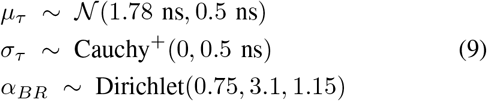

which reflect the priors in Eqs. (2) and (3).

Note that a basic assumption of Bayesian hierarchical models is that different VOI are exchangeable, i.e. the joint distribution 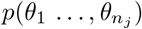 of the group-level parameters *θ*_*j*_ is invariant under permutation of the indices *j*. In our case, exchangeability is ensured by sampling independently from

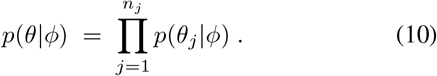

We allow also for partial exchangeability. In some cases, we have prior knowledge and we can form subgroups of VOI and each subgroup has its own exchangeable hierarchical model with a common population distribution. More specifically, let us assume that the first *s*_*j*_ VOI belong to a subgroup, then the permutation invariance applies to the sets of indices *j* = 1, …, *s*_*j*_ and *j* = *s*_*j*_ +1, …, *n*_*j*_ separately. The joint posterior distribution for *θ* and *ϕ* is

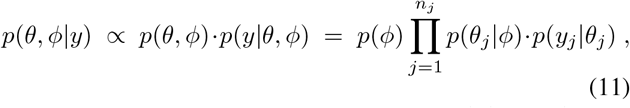

where the priors *p*(*ϕ*) are given in Eq. (9) and the conditional distribution *p*(*θ*_*j*_ |*ϕ*) is simply a short-hand notation for the distributions in Eq. (8). Note that *p*(*θ, ϕ* | *y*) does not factorize.

In the present study, we construct hierarchical models for VOI of subject S3 containing venous and arterial blood. The working hypothesis is that the lower oxygen content of venous blood compared to arterial blood is reflected through different oPs lifetimes. We set up four different hierarchical models, as shown schematically in Fig. 3. In model *m*_0_ the oPs lifetime is sampled from a common distribution for all six VOI under consideration (aorta, left atrium and ventricle, axillary vein, right atrium and ventricle). For model *m*_1_ we separate two subgroups of three VOI each according to their blood content. This means that the three oPs lifetimes 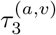and branching ratios 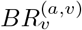 of the VOI inside one subgroup are sampled from a common population distribution. This population distribution is different for the arterial (index *a*) and venous blood (index *v*) subgroups. Lastly, in model 2 and 3 we combine the heart chambers and compare them with the axillary vein or aorta. For these two models, the fit data encompasses only three TDD.

**Fig. 3:**
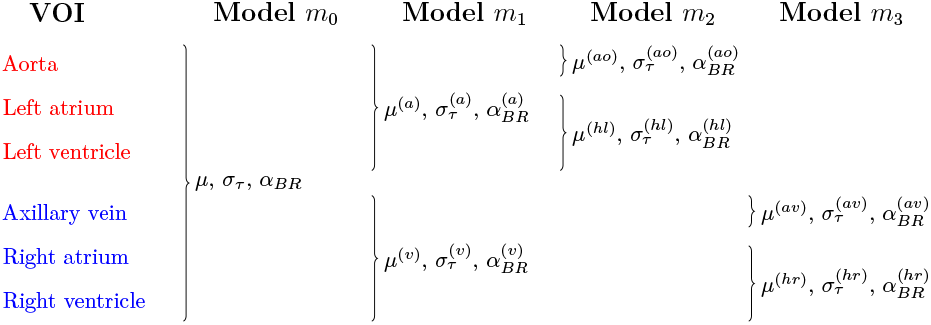
Hierarchical models for the comparison of oPs lifetimes in VOI with venous (blue) or arterial blood (red).

The purpose is to compare the performance of the different hierarchical models, even if the number of fitting parameters is different. We evaluate the hierarchical models in Fig. 3 according to their out-of-sample predictive accuracy while taking into account the uncertainty of its estimate. LOO-CV in combination with a Pareto smoothed importance sampling is a robust and efficient approach for estimating the out-of-sample predictive accuracy without refitting the model (see Ref. [41], [42]).

The basic idea is find an approximation for the the expected log point-wise predictive density (elpd) for *i* = 1, …, *ñ* new data points 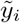 given the data generating model *m* and the data *y*. For a hierarchical model *m*, the posterior predictive density 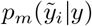 for new data is the marginalized distribution

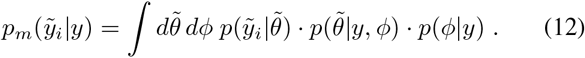

where 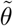 are new group-level parameters for the data 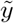. In the absence of independent validation data, we can use LOO-CV to estimate the elpd

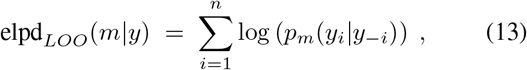

where *y*_−*i*_ are all observations in *y* except *y*_*i*_ and predictive density *p*_*m*_(*y*_*i*_ | *y*_−*i*_) is analogous to Eq. (12). In order to compare two different models *m*_*a*_ and *m*_*b*_ from Fig. 3, we can simply evaluate the sign of the eldp difference

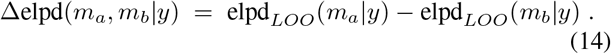

## Results

First, we present in Fig. 4 the histoimages of all detected 3*γ*E of the three subjects. For each count in Fig. 4 we are able to determine a time difference between the prompt and the annihilation photons. The voxel size is chosen according to the CT image, i.e. 0.98 × 0.98 × 3.0 mm for S1, 1.52 × 1.52 × 3.0 mm^3^ for S2 and 1.52 × 1.52 × 1.65 mm^3^ for S3. These histoimages resemble typical distribution of the radiopharmaceuticals administered to the three subjects, i.e. there is a prominent uptake of [^68^Ga]Ga-DOTA-TOC in S1’s meningioma and spleen, the clearly visible parotid glands of S2 and the well perfused heart, kidneys and right veins through which the ^82^Rb was injected into S3. This, of course, correlates well with the coincidence PET images shown in Fig. 1.

**Fig. 4:**
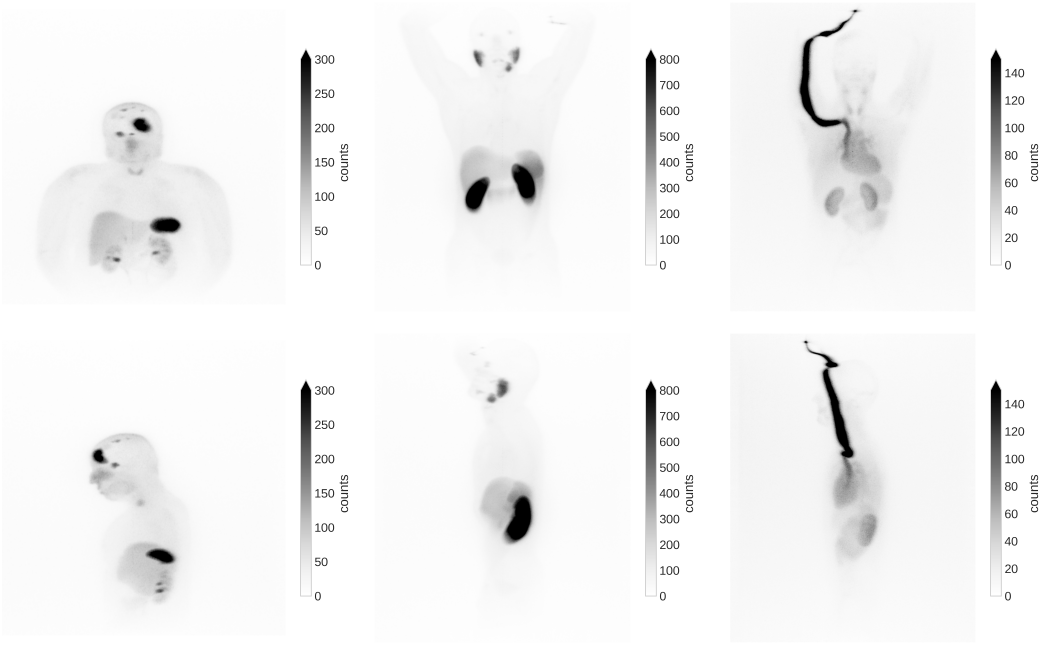
Coronal (top row) and sagittal (bottom row) MIP of the histoimages for the subjects S1 (left), S2 (center) and S3 (right). These MIP of the histoimages resemble typical coincidence PET images for these tracers. Note that the histoimages include all detected 3*γ*E, i.e. also the random events.

The main results of this study are the TDDs of selected VOIs that are shown in Figs. 5, 6 and 7 for the three subjects. The error bars shown in these TDD are the relative standard deviation of the background regions with Δ*t* < − 2 ns. The noise level for different organs varies substantially. The relative error in the background region ranges from 0.2 % for S2’s liver to 4.6 % for S3’s thyroid.

**Fig. 5:**
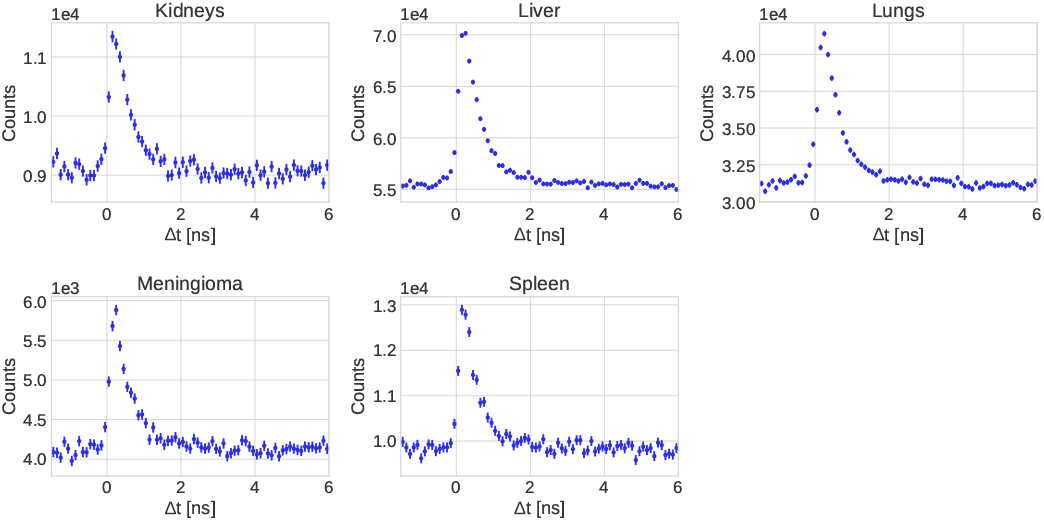
Measured TDD for selected VOI from subject S1.

**Fig. 6:**
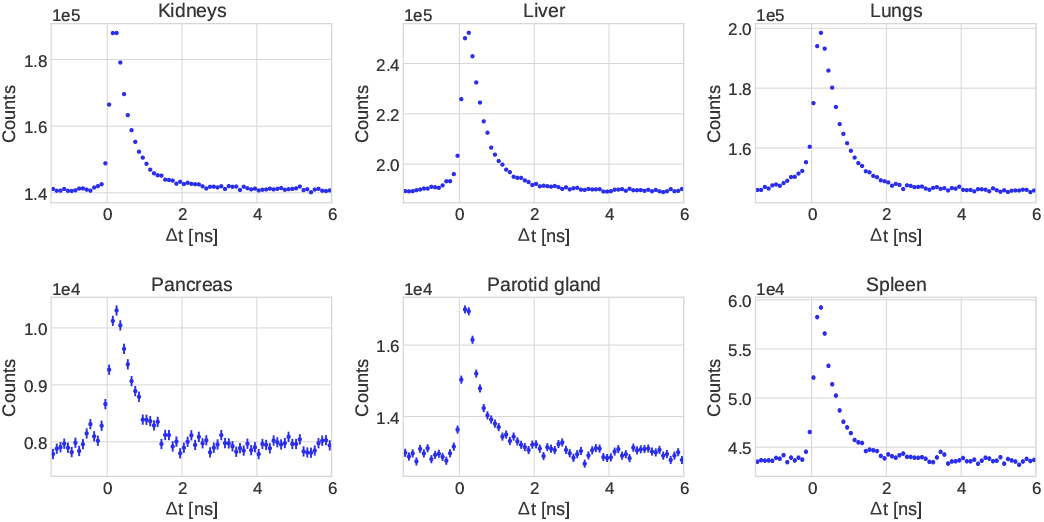
Measured TDD for selected VOI from subject S2.

**Fig. 7:**
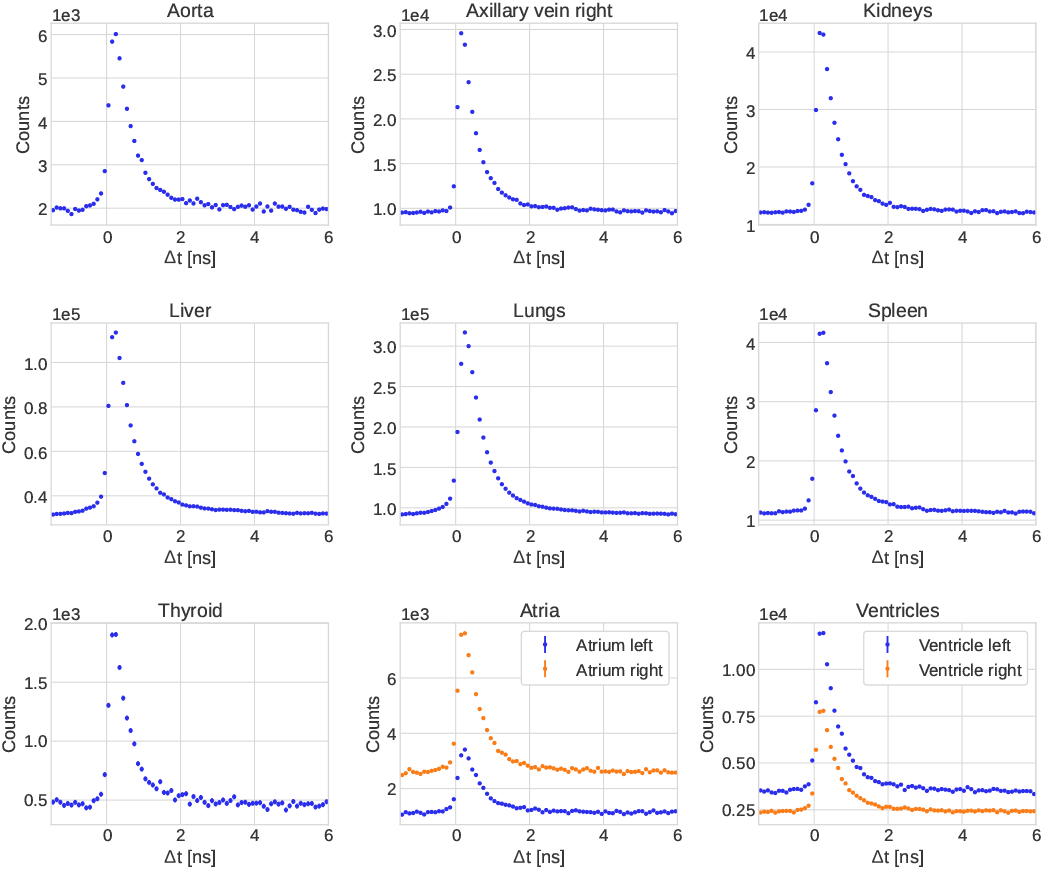
Measured TDD for selected VOI from subject S3.

When comparing selected organs across the three subjects, the count statistics for S1 and S2 are much higher than for S3 (see the MIP in Fig. 4). However, the peak signal-to-background ratio (pSBR) is significantly higher for S3. Despite having low statistical noise, as seen e.g. in the liver of S1 and S2, the pSBR does not go beyond ≈ 1.3. On the other hand, for S3 we found that pSBR is consistently above ≈ 3.3 for the VOI under consideration.

From the measured TDD we can determine the oPs lifetime through the Bayesian fitting procedure described in Sec. II-C. For the VOI with decent count statistics and SBR, Tabs. III, IV and V report the fitted oPs lifetimes and branching ratios of the three fit components (pPs, direct annihilation and oPs). The HDI are taken at 68%. Note that the posterior distribution of *τ*_3_ is mostly bell-shaped and it would therefore also make sense to estimate the posterior variance through the common point estimate. However, the posterior distributions of *BR*_*c*_ are far from being bell-shaped. Estimating a variance would be meaningless. As expected, the high pSBR leads to considerable uncertainty on *τ*_3_ for S1. While the first moment of *τ*_3_’s posterior distribution shifts compared to the prior in Eq. (2), the HDI is only slightly smaller than the prior’s. Except for the pancreas, the VOI of S2 give already a reasonable constrain on *τ*_3_. In line with expectations, Tab. V shows that S3’s TDD are most constraining on the posterior distributions of *τ*_3_ and *BR*_*c*_.

The fit predictions are visualized in Fig. 8. For the most interesting VOI we show the measured TDD together with the posterior mean for the three fit components and their sum. Note that the fit ranges from − 1.5 ns to 8 ns. The limit of 6 ns in Fig. 8 is selected purely for improved visualization.

**Fig. 8:**
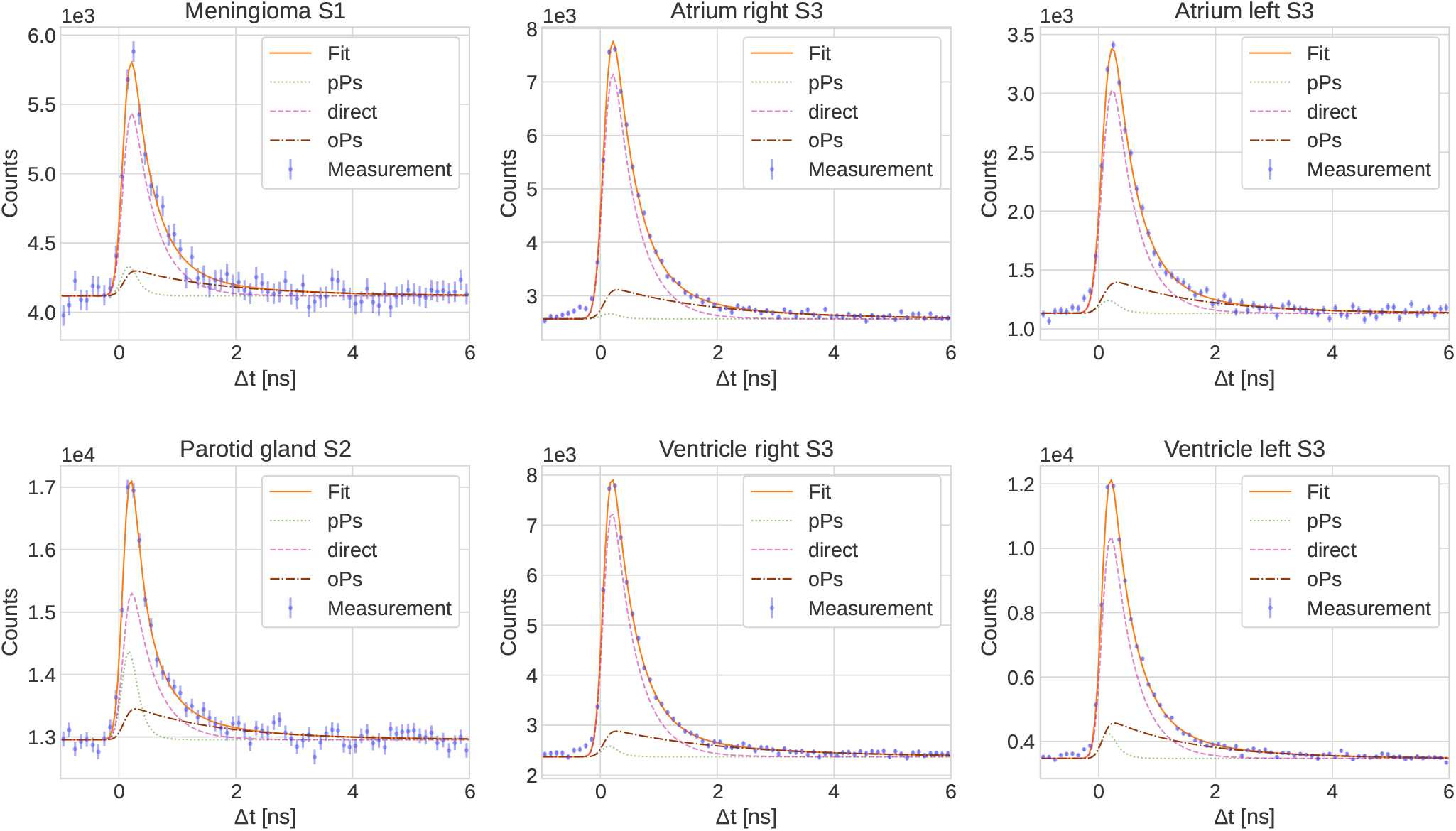
TDD and fit prediction (posterior mean) for selected VOI.

The hierarchical models from Fig. 3 are fitted to the aorta, the right axillary vein, the ventricles and the atria of S3 and we report the results in Tab. VI. The posterior distribution for *σ*_*τ*_ is strongly skewed and truncated at 0. Therefore, we only quote the 68% HDI for these hyperparameters.

The 68% 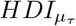 of 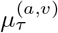 of *m*_1_ in Tab. VI overlap slightly, but the expectation value of the arterial blood VOI 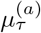 is almost 0.4 ns lower than for the arterial blood. If we were to fix both *τ*_1,2_ in the hierarchical model fit, the two 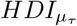 would not overlap, i.e. 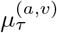 would tend to lower values and the 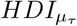 would be [1.10, 1.45] ns and [1.49, 1.81]. The HDI for both 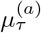 and 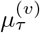 are significantly smaller than the HDI of the prior distributions in Eq. (9), meaning that the measured TDD are strongly constraining the hyperparameters. Fig. 9 shows the posterior distribution of the hyperparameters in the models *m*_0,1_. When comparing the elpd_*LOO*_ of the two models *m*_0,1_, we notice only a slight improvement for *m*_1_, i.e. Δelpd(*m*_0_, *m*_1_| *y*) < 0. The difference is, however, much smaller than the estimated standard error of elpd_*LOO*_ quoted in the last column of Tab. VI. When comparing the hyperparameters of model *m*_2_ in Tab. VI, it seems that 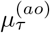 approaches higher values (similar to water), while the left heart chambers with oxygenated blood seem to drive the oPs lifetime to smaller values. In model *m*_3_ however, *τ*_3_ in the right axillary vein is in line with the one in the right heart chambers.

**Fig. 9:**
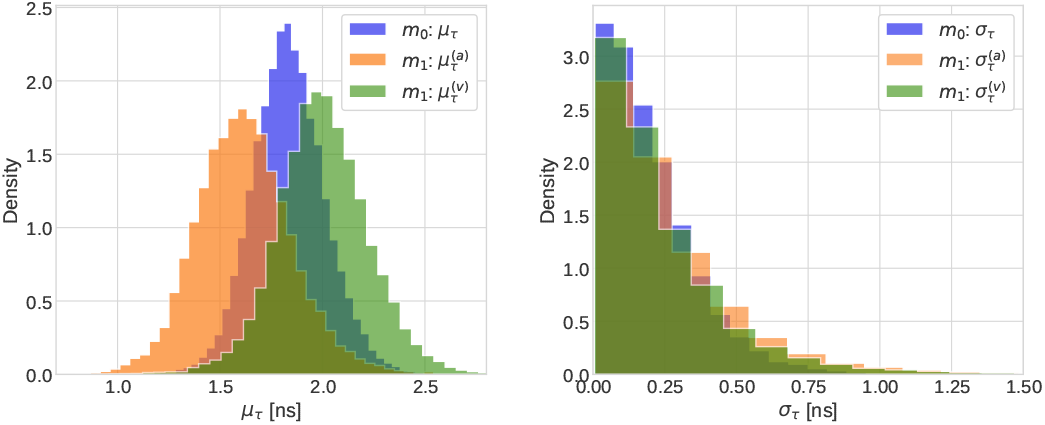
Comparison between the sampled posterior distributions of the hyperparameters of the models *m*_0_ and *m*_1_ (see Fig. 3).

## IV. Discussion

This study is the first to use a commercial PET/CT scanner and the first to use a intravenous administration of the PET tracer to measure the oPs lifetime in vivo. This comprised the selection of 3*γ*E and the determination the time difference between the prompt and annihilation photon is possible on a commercial medical device. We hope that our results can brake the ground for further investigations and developments of PALS in medical applications.

A first in vivo oPs lifetime measurement was previously presented in Ref. [22]. The used scanner is a prototype scanner with a standard FOV that is not commercially available and the tracer used of oPS lifetime measurement was administered into the operating field, which is not translatable to routine clinical examinations. Simulations in Ref. [43] suggested that the sensitivity of Quadra is about 400 times higher than J-PET’s. Figs. 5, 6 and 7 do not suggest such a large difference between the two scanners, but there is no denial that Quadra outperforms the current version of the J-PET scanner with respect to count statistics.

However, also in the present study with the commercial Quadra PET/CT, the count statistics of 3*γ*E is the main issue. The pSBR is significantly lower for the ^68^Ga-based compounds compared to ^82^Rb and the background noise is typically higher. The lower prompt photon branching ratio of ^68^Ga and the longer scan times for S1 and S2 increase random 3*γ*E and therefore the background in the measured TDD. On the other hand, [^82^Rb]Cl has only a marginal uptake in any tissue due to the large decay constant. This means that oPs lifetime in S3 is likely to reflect the properties of blood and not of the tissue that takes up the tracer.

The fitting function in Eq. (1) inherently induces strong correlations, in particular among the Gaussian parameters *σ* and Δ as well as among each *τ*_*i*_ and the corresponding *BR*_*c*_ (see the pair plot in Fig. 15 of Ref. [35]). Having a low noise and background level is therefore crucial for disentangling these correlations, which helps limiting the uncertainties in the marginalized posterior distributions. We chose a rather conservative approach to the TDD fitting procedure. From the parameters in Eq. (1), we only fix the TDD’s background *b*. Through our event selection procedure, the TDD contains random 3*γ*E with negative time differences. As shown in Ref. [35] with ^18^F measurements, the crystal distance threshold in the event selection gives a flat background throughout positive and negative time differences. We are therefore confident that estimating *b* from events with negative time differences yields an accurate estimate. Note that allowing for *b* to be a fit parameter would introduce a strong correlation with *τ*_3_ since oPs is dominant in tail region of the TDD (starting at around 2 ns). For the background estimation, Ref. [22] takes a similar approach. The priors on *τ*_1,2_ in Eq. (2) are quite narrow, reflecting our strong prior knowledge on these lifetimes and their independence from the surrounding material. Despite this strong prior we allow for a variation of *τ*_1,2_. As shown e.g. in Ref. [37] measurements of *τ*_1,2_ do show some variation across different measurements. The rather uninformative prior on *τ*_3_ expresses our lack of prior knowledge about oPs lifetimes in tissue. The same applies to the priors on *BR*_*c*_.

Other research groups have made approximations to reduce the uncertainty on *τ*_3_ whenever the TDD’s count statistics is low. In Ref. [38] the J-PET collaboration fixes the pPs lifetime *τ*_1_ and direct annihilation time *τ*_2_. Ref. [34] relies on a simplified two-lifetime-components model, i.e. one short- and one long-lived. The authors of Ref. [22] go even further by fixing the branching ratios of a two-component model, effectively leaving only *τ*_3_ and a scale parameter as free parameters. These approaches could limit the interpretability of the longer lived component as being oPs in a physical sense and may therefore limit the possibility to compare with in vitro PALS measurements.

In general, the oPs lifetimes shown in Tabs. III, IV and V are of the same order of magnitude as in water. At 37^*°*^ water temperature, Ref. [37] reports *τ*_3_ ≈ 1.78 ns. In general, it is fair to say that at this point we lack an interpretation for the fitted *τ*_3_ values. Only future large scale in vitro, preclinical and clinical PALS measurements will eventually allow for a rigorous interpretation of *τ*_3_ as a biomarker. In this vein, our study is merely showing what commercial PET systems are capable of and what the current challenges for in vivo PALS measurements are.

The *τ*_3_ values of S1 in Tab. III are affected by a rather large 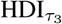. This is a consequence of the low pSBR. In contrast, the meningioma VOI of S1 is promising. A Gd-enhanced MR scan prior to the PET scan showed a good perfusion of the meningioma and also the coincidence PET image does not show any larger areas without uptake. A *τ*_3_-increasing hypoxia is therefore not expected, which is in line with the lower expectation value oPs lifetime in Tab. III.

**TABLE III:** Fit results for selected organs from S1.

| VOI | $\tau_3$ [ns] | HDI$_{\tau_3}$ [ns] | $BR_1$ | HDI$_{BR_1}$ | $BR_2$ | HDI$_{BR_2}$ | $BR_3$ | HDI$_{BR_3}$ |
| --- | --- | --- | --- | --- | --- | --- | --- | --- |
| Kidneys | 1.65 | [1.12, 2.2] | 0.08 | [0.0, 0.06] | 0.71 | [0.68, 0.89] | 0.21 | [0.10, 0.26] |
| Liver | 1.92 | [1.52, 2.28] | 0.02 | [0.0, 0.02] | 0.73 | [0.71, 0.78] | 0.25 | [0.22, 0.28] |
| Lungs | 1.98 | [1.62, 2.28] | 0.07 | [0.0, 0.09] | 0.66 | [0.61, 0.74] | 0.28 | [0.24, 0.3] |
| Meningioma | 1.58 | [1.10, 1.94] | 0.06 | [0.0, 0.08] | 0.67 | [0.60, 0.78] | 0.27 | [0.20, 0.32] |
| Spleen | 2.29 | [1.89, 2.66] | 0.11 | [0.05, 0.16] | 0.59 | [0.51, 0.68] | 0.30 | [0.26, 0.34] |

**TABLE IV:** Fit results for selected organs from S2.

| VOI | $\tau_3$ [ns] | HDI$_{\tau_3}$ [ns] | $BR_1$ | HDI$_{BR_1}$ | $BR_2$ | HDI$_{BR_2}$ | $BR_3$ | HDI$_{BR_3}$ |
| --- | --- | --- | --- | --- | --- | --- | --- | --- |
| Kidneys | 1.70 | [1.50, 1.86] | 0.11 | [0.08, 0.13] | 0.63 | [0.6, 0.66] | 0.26 | [0.24, 0.28] |
| Liver | 1.46 | [1.16, 1.68] | 0.04 | [0.0, 0.05] | 0.7 | [0.67, 0.77] | 0.26 | [0.22, 0.28] |
| Lungs | 1.35 | [1.00, 1.57] | 0.05 | [0.0, 0.06] | 0.61 | [0.56, 0.72] | 0.34 | [0.27, 0.38] |
| Pancreas | 1.59 | [1.07, 2.06] | 0.05 | [0.0, 0.06] | 0.77 | [0.71, 0.88] | 0.18 | [0.11, 0.24] |
| Parotid gland | 1.49 | [1.04, 1.76] | 0.18 | [0.13, 0.24] | 0.52 | [0.42, 0.64] | 0.30 | [0.23, 0.35] |
| Spleen | 1.63 | [1.28, 1.89] | 0.07 | [0.03, 0.1] | 0.69 | [0.64, 0.76] | 0.24 | [0.21, 0.27] |

Since [^82^Rb]Cl is a perfusion tracer, we expect the oPs lifetimes listed in Tab. V to be mostly driven by the molecular structure (i.e. oxygenation) of the blood’s constituents. The meta-analysis in Ref. [44] found an average difference of 4.92 ± 0.61 kPa in partial pressure of oxygen *P*_*a,v*_*O*_2_ between venous and arterial blood. In conjunction with the results of Ref. [15], there is the potential to measure *P*_*a,v*_*O*_2_ through oPs lifetime measured in S3. Considering the VOI in Tab. V that contain arterial or venous blood, the expectation values for *τ*_3_ seem to be consistently lower for arterial blood. However, the 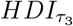 of the left and right heart chambers overlap, making these differences non-significant.

**TABLE V:** Fit results for selected organs from S3.

| VOI | $\tau_3$ [ns] | HDI$_{\tau_3}$ [ns] | $BR_1$ | HDI$_{BR_1}$ | $BR_2$ | HDI$_{BR_2}$ | $BR_3$ | HDI$_{BR_3}$ |
| --- | --- | --- | --- | --- | --- | --- | --- | --- |
| Aorta | 1.60 | [1.34, 1.80] | 0.02 | [0.0, 0.02] | 0.67 | [0.65, 0.72] | 0.31 | [0.28, 0.33] |
| Axillary vein | 1.80 | [1.66, 1.94] | 0.09 | [0.08, 0.1] | 0.66 | [0.64, 0.68] | 0.26 | [0.24, 0.27] |
| Kidneys | 1.72 | [1.55, 1.86] | 0.04 | [0.02, 0.06] | 0.70 | [0.68, 0.72] | 0.26 | [0.25, 0.28] |
| Liver | 1.44 | [1.25, 1.58] | 0.01 | [0.0, 0.01] | 0.64 | [0.61, 0.68] | 0.35 | [0.32, 0.38] |
| Lungs | 1.52 | [1.32, 1.66] | 0.02 | [0.0, 0.03] | 0.67 | [0.65, 0.71] | 0.31 | [0.28, 0.33] |
| Spleen | 1.66 | [1.48, 1.80] | 0.03 | [0.0, 0.03] | 0.69 | [0.66, 0.72] | 0.29 | [0.27, 0.30] |
| Thyroid | 1.83 | [1.51, 2.08] | 0.04 | [0.0, 0.05] | 0.69 | [0.65, 0.74] | 0.28 | [0.24, 0.30] |
| Atrium left | 1.50 | [1.15, 1.72] | 0.02 | [0.0, 0.03] | 0.71 | [0.68, 0.77] | 0.26 | [0.22, 0.29] |
| Atrium right | 1.76 | [1.46, 1.99] | 0.01 | [0.0, 0.01] | 0.72 | [0.7, 0.75] | 0.27 | [0.24, 0.29] |
| Ventricle left | 1.44 | [1.22, 1.60] | 0.04 | [0.02, 0.07] | 0.67 | [0.63, 0.72] | 0.29 | [0.26, 0.31] |
| Ventricle right | 1.96 | [1.69, 2.18] | 0.02 | [0.0, 0.02] | 0.72 | [0.7, 0.75] | 0.26 | [0.24, 0.28] |

Given the Bayesian framework that is used, it is feasible to go beyond a mere comparison of single VOI fit values to investigate the oPs lifetime in the heart chambers of S3 by using a hierarchical model. As was the case for the single VOI fits in Tab. V, the hyperparameters’ 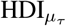 of the posterior distributions are smaller compared to the priors. However, 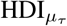 in Tab. VI is comparable to 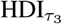 in Tab. V. This indicates that is is feasible to constrain 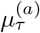 and 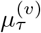 in *m*_1_, i.e. assuming that the oxygenation and oPs lifetime is different in venous and arterial blood. However, the two models *m*_0_ and *m*_1_ only marginally differ in their elpd and therefore perform very similarly on the available data, but still indicate that the model assuming a difference in oPs lifetime between arterial and venous blood is better Δelpd(*m*_0_, *m*_1_ | *y*) < 0. Even fixing some of the fit parameters does not significantly improve the difference of 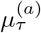and 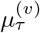. This might indicate insufficient count statistics in the available data. Adapting an approach to the *τ*_3_ fitting in S3 that is more in line with Refs. [22], [38] might allow for a stronger statement. We leave this question open for future studies to answer.

**TABLE VI:** Fit results for the hyperparameters in the hierarchical models from Fig. 3 together with the LOO estimate of elpd_*LOO*_ and its standard error.

| Model | $\mu_\tau$ | $\text{HDI}_{\mu_\tau}$ | $\text{HDI}_\sigma$ | $BR_3$ | $\text{HDI}_{BR_3}$ | $\text{elpd}_{\text{LOO}}$ |
| --- | --- | --- | --- | --- | --- | --- |
| $m_0$ | 1.84 | [1.66, 2.0] | [0.01, 0.25] | 0.33 | [0.22, 0.42] | $-3211.73 \pm 36.62$ |
| $m_1^{(a)}$ | 1.62 | [1.4, 1.84] | [0.01, 0.3] | 0.32 | [0.18, 0.43] | $-3211.95 \pm 36.40$ |
| $m_1^{(v)}$ | 2.0 | [1.78, 2.21] | [0.0, 0.27] | 0.3 | [0.17, 0.4] | |
| $m_2^{(ao)}$ | 1.75 | [1.41, 2.07] | [0.0, 0.51] | 0.29 | [0.1, 0.41] | $-1556.17 \pm 23.10$ |
| $m_2^{(hl)}$ | 1.48 | [1.2, 1.68] | [0.01, 0.33] | 0.3 | [0.15, 0.42] | |
| $m_3^{(av)}$ | 1.86 | [1.59, 2.14] | [0.0, 0.47] | 0.26 | [0.08, 0.35] | $-1640.49 \pm 22.03$ |
| $m_3^{(hr)}$ | 1.85 | [1.54, 2.13] | [0.01, 0.46] | 0.31 | [0.14, 0.41] | |

We refrain from determining *τ*_3_ at the voxel level for the three subjects in our study. Such an oPs lifetime image is not feasible with the current level of count statistics, with our fitting methodology and a reasonably small voxel size.

## V. Conclusions

In this study, the feasibility in vivo oPs lifetime measurements using a commercial LAFOV PET/CT and a intravenous tracer administration was shown. A Bayesian framework was developed to enable a conservative approach for extracting the lifetime and to more accurately determine the statistical errors in the fitting procedure. With [^82^Rb]Cl, organ-level oPs lifetime can be determined with reasonable statistical uncertainty, though SBR remains a challenge, particularly for ^68^Ga and voxel-level *τ*_3_ measurements. Future validation studies are warranted to assess the potential for measuring the oxygenation level and to investigate the extension of voxelwise estimation of oPs lifetime.

## Data Availability

Evaluated data from the 82Rb dataset is available in the Zenodo repository https://doi.org/10.5281/zenodo.11243763. The remaining datasets generated and/or analysed during the current study are available from the corresponding author on reasonable request.

